# Cystic fibrosis risk variants confer protection against inflammatory bowel disease

**DOI:** 10.1101/2024.12.02.24318364

**Authors:** Mingrui Yu, Qian Zhang, Kai Yuan, Aleksejs Sazonovs, Christine Stevens, Laura Fachal, the International Inflammatory Bowel Disease Genetics Consortium, Carl A. Anderson, Mark J. Daly, Hailiang Huang

## Abstract

Genetic mutations that yield defective cystic fibrosis transmembrane regulator (*CFTR*) protein cause cystic fibrosis, a life-limiting autosomal recessive Mendelian disorder. A protective role of *CFTR* loss-of-function mutations in inflammatory bowel disease (IBD) has been suggested, but its evidence has been inconclusive and contradictory. Here, leveraging the largest IBD exome sequencing dataset to date, comprising 38,558 cases and 66,945 controls in the discovery stage, and 35,797 cases and 179,942 controls in the replication stage, we established a protective role of CF-risk variants against IBD based on evidence from the association test of *CFTR* delF508 (p-value=8.96E-11) and the gene-based burden test of CF-risk variants (p-value=3.9E-07). Furthermore, we assessed variant prioritization methods, including AlphaMissense, using clinically annotated CF-risk variants as the gold standard. Our findings highlight the critical and unmet need for effective variant prioritization in gene-based burden tests.

---

Genetic mutations that yield defective cystic fibrosis transmembrane regulator (CFTR) protein are known to cause cystic fibrosis (CF)^1^, one of the most common life-threatening autosomal recessive genetic disorders among individuals of European ancestry. *CFTR* is a transmembrane chloride ion channel gene highly expressed in epithelial cells^2^. Defective *CFTR* protein located in the epithelial cell membrane results in defective chloride ion transport^3^, which leads to water depletion at the cell surface and thick mucus buildup in organs such as the lung, pancreas, sweat gland, and gut. Cellular signaling pathways through which the innate immune system elicits proper immune responses are dysregulated in CF^4–10^, leading to hyperinflammation. However, it is unclear whether *CFTR* mutations are intrinsically pro-inflammatory or whether the heightened inflammation observed in CF results from a heightened response to exacerbated pathogen exposure^11–14^.

Inflammatory bowel disease (IBD) is a disorder with chronic inflammation in the gastrointestinal tract. Although earlier research suggested that loss-of-function variants in the *CFTR* gene may lead to a lower prevalence of intestinal inflammation^15^ and potentially protect against IBD, evidence has been inconsistent with studies showing conflicting results - some linking these variants to increased risk, others suggesting protection, and some finding no effect at all^16–19^. Here, leveraging the largest IBD exome sequencing dataset to date, comprising 38,558 cases and 66,945 controls in the discovery stage, and 35,797 cases and 179,942 controls in the replication stage, we provide conclusive evidence that CF-risk variants in *CFTR* confer a protective effect against IBD, and demonstrated the importance of variant prioritization in gene-based burden tests.

## RESULTS

### Study subjects

#### Broad Institute (Discovery)

Exome sequencing was performed at the Broad Institute. Study subjects were recruited from different centers and shared with the International Inflammatory Bowel Disease Genetics Consortium (IIBDGC) for 38,558 IBD cases and 66,945 controls after quality control analysis (Table 1). Among the cases, 21,478 have Crohn’s disease (CD), 14,353 have ulcerative colitis (UC), and 2,727 have IBD unclassified (IBD-U). *Sanger Institute (Replication)*: Additional exome and whole genome sequencing were undertaken at the Sanger Institute. Whole exome sequencing was performed on 10,722 CD, 13,147 UC, and 6,211 IBD-U, which were matched with whole exome sequencing data obtained from 168,100 controls from the UK Biobank. Extensive quality control was undertaken to harmonize the case and control exome sequencing data (Methods). Whole genome sequencing was performed on another independent sample of 5,717 CD and 11,842 controls. Details on sequencing and data quality-control protocols are described in Methods.

**Table 1.** Study sample sizes. The number of individuals of European ancestry included in the association analyses, post-quality control (Methods). WES: whole-exome sequencing; WGS: whole-genome sequencing.

| Center | Stage | Method | Controls | CD | UC | IBD-U |
| --- | --- | --- | --- | --- | --- | --- |
| Broad | Discovery | WES | 66,945 | 21,478 | 14,353 | 2,727 |
| Sanger | Replication | WES | 168,100 | 10,722 | 13,147 | 6,211 |
|  |  | WGS | 11,842 | 5,717 | 0 | 0 |

### CF-causing variant deltaF508 confers a protective effect against IBD

The *CFTR* deltaF508 is an in-frame deletion variant commonly observed in the European population, with a minor allele frequency (MAF) of 1.5% in non-Finnish Europeans (chr7:117559590:ATCT:A in Genome Reference Consortium Human Build 38 [GRCh38]). Homozygous deltaF508 has full penetrance for CF and is the most common CF-causing variant. Single variant association analysis in the Broad discovery dataset using a logistic mixed model (Methods) found that deltaF508 had a significant protective effect against both CD (p-value=1.53E-7, OR=0.73 [0.65-0.82, 95% CI]) and UC (p-value=3.35E-4, OR=0.79 [0.69-0.90, 95% CI], Table 2). This association also reached nominal significance for CD in the Sanger WES and WGS datasets (p-value=3.52E-02 and 8.32E-3, Table 2) with both protective effects. After meta-analysis, the protective effect of deltaF508 on CD reached genome-wide significance (p-value=5.5E-9). There are no known IBD-associated variants (defined as variants with posterior inclusion probability > 5% from fine-mapping^20^ or reported in the published sequencing and genome-wide association studies^21–23^) within 1 million base pairs of deltaF508, therefore it is highly unlikely that the deltaF508 association tags a known IBD genetic association.

**Table 2.** Association between deltaF508 and CD, UC, and IBD. MAF: minor allele frequency; BETA, SE: effect size, and standard error from a logistic mixed model; OR (95% CI): 95% confidence interval of odds ratio (OR).

| Study | Subtype | MAF (case) | MAF(control) | p-value | BETA | SE | OR (95% CI) |
| --- | --- | --- | --- | --- | --- | --- | --- |
| Broad WES<br>(Discovery) | CD | 0.0097 | 0.0136 | 1.53E-07 | -0.305 | 0.059 | 0.65 - 0.82 |
|  | UC | 0.0096 | 0.0136 | 3.35E-04 | -0.234 | 0.066 | 0.69 - 0.90 |
|  | IBD | 0.0097 | 0.0136 | 8.50E-10 | -0.289 | 0.047 | 0.68 - 0.82 |
| Sanger WES<br>(Replication) | CD | 0.0135 | 0.0161 | 3.52E-02 | -0.136 | 0.064 | 0.77 - 0.99 |
|  | UC | 0.0141 | 0.0161 | 2.04E-01 | -0.073 | 0.058 | 0.83 - 1.04 |
|  | IBD | 0.0137 | 0.0161 | 1.47E-02 | -0.108 | 0.044 | 0.82 - 0.98 |
| Sanger WGS<br>(Replication) | CD/IBD | 0.0114 | 0.0157 | 8.32E-03 | -0.310 | 0.117 | 0.58 - 0.92 |
| **Meta-analysis** | **CD** | - | - | **5.49E-09** | **-0.238** | **0.041** | **0.73 - 0.85** |
|  | **UC** | - | - | **9.95E-04** | **-0.143** | **0.043** | **0.79 - 0.94** |
|  | **IBD** | - | - | **8.96E-11** | **-0.201** | **0.031** | **0.77 - 0.87** |

To limit the potential for confounded CF status to bias the allele frequency estimates in our cases and controls, we repeated the association analysis after excluding 4 cases and 47 controls who were homozygous carriers of deltaF508 in the discovery and replication cohorts combined, as well as 37 cases and 170 controls deemed potential compound heterozygous carriers (Supplementary Tables 1), defined as individuals with two or more variants annotated as “CF-causing” or “Varying clinical consequence” in the Clinical and Functional Translation of *CFTR*^*24*^ (CFTR2) database. We found that the associations between *CFTR* deltaF508 and CD/UC/IBD remain significant (Supplementary Tables 2), supporting the conclusion that deltaF508 plays a protective role in IBD.

### *CFTR* mutations confer protection against IBD in gene-based burden tests

The CFTR2 database contains a comprehensive list of *CFTR* variants known to impair *CFTR* protein function and cause CF. The Broad discovery dataset captured 1036 *CFTR* variants (Supplementary Tables 3). 170 of the 1035 CFTR variants were annotated in CFTR2 as “CF-causing” (109), “Varying clinical consequence” (33), “Non CF-causing” (19), or “Unknown significance” (9). The remaining 865 variants did not have an annotation in CFTR2. We defined “CF-risk” variants (142) as variants annotated as “CF-causing” or “Varying clinical consequences” in the CFTR2 database, and “Non-CF-risk” variants (893) as variants annotated as “Non CF-causing”, “Variants of unknown significance” in CFTR2, or without annotation in CFTR2.

While no variants other than deltaF508 reached even nominal levels of significance (p-value<0.01) in our single variant tests, CF-risk variants are predominantly protective against IBD (p-value=0.002, binomial test using variants with minor allele count ≥ 10 in the Broad dataset, Figure 1a). Conversely, non-CF-risk variants were not enriched with protective effects (p-value=0.35, binomial test using variants with minor allele count ≥ 10 in the Broad dataset, Figure 1b).

**Figure 1:**
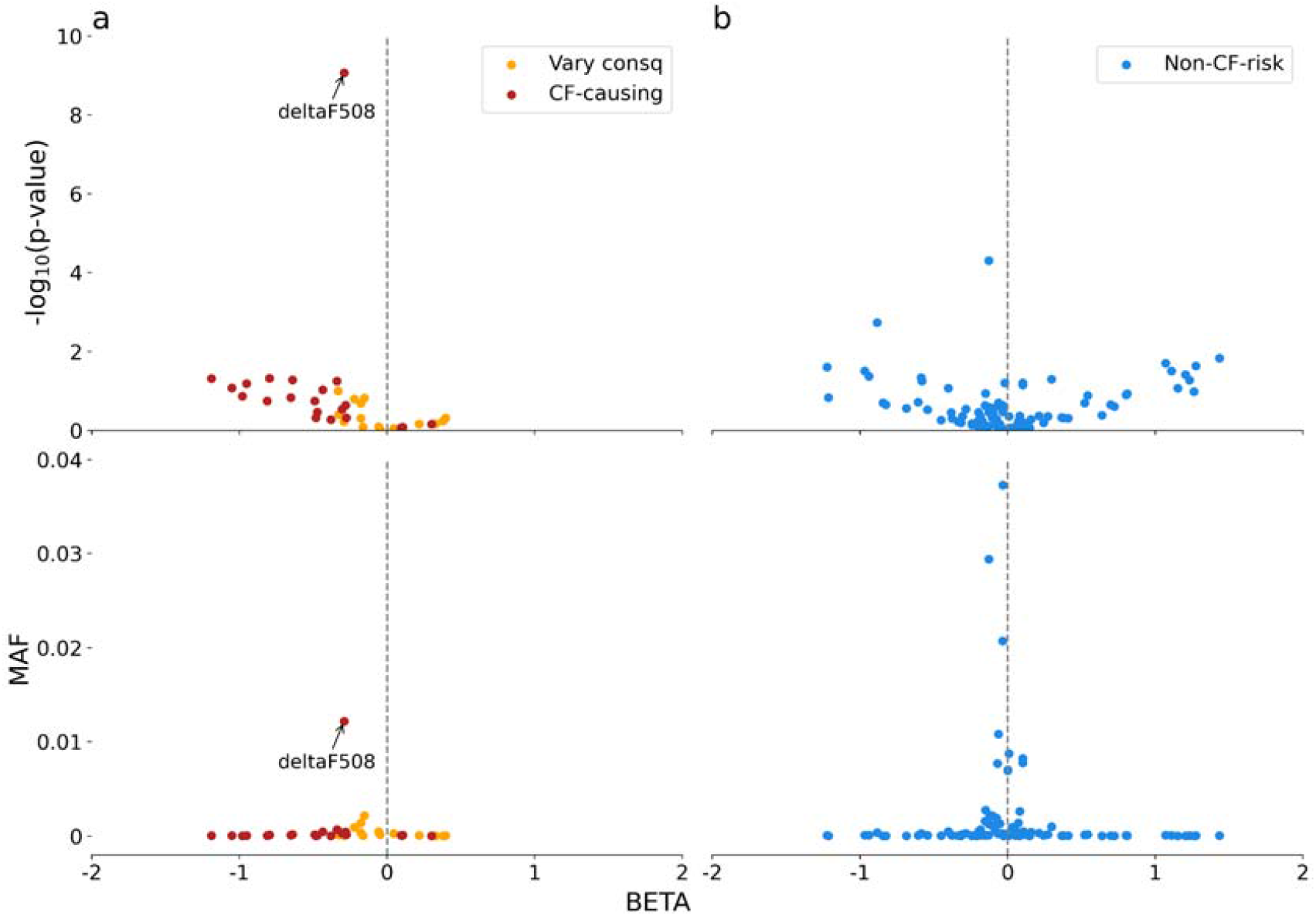
Single-variant association tests for CFTR variants with minor allele count (MAC) ≥ 10 in the Broad dataset. **a)** Distribution of p-values (from logistic mixed model) and MAF for 37 CF-risk variants. **b)** Distribution of p-values (from logistic mixed model) and MAF for 96 non-CF-risk variants.

Based on these observations, we hypothesize that variants impairing *CFTR* function are protective against IBD. To test this hypothesis, we did an unweighted burden test of *CFTR* using the 141 CF-risk variants (deltaF508 not included as itself confers a significant protective effect). We found these variants in aggregation have a significant protective effect on IBD in the Broad dataset (P=4.3E-06, beta=-0.21, se=0.047) (Table 3), which was replicated in the Sanger WES dataset at nominal significance (P=0.037, beta=-0.11, se=0.052) and in the Sanger WGS dataset for the direction and effect size with CD (p-value=0.073, beta=-0.15, se=0.088). The Sanger WGS is much smaller in the sample size thus, the lack of significance is likely due to its limited power. Combining all three datasets using the fixed-effect meta-analysis, we found CF-risk variants in *CFTR* collectively confer a protective effect of -0.164 (OR=0.85) with p-value=3.9E-7. This association was significant when tested for CD and UC separately (meta-analysis for CD: beta=-0.13, se=0.039, p-value=8.8E-04; UC: beta=-0.23, se=0.049, p-value=3.6E-06) (Supplementary Table 4). On the contrary, we did not observe a significant association with IBD using the non-synonymous variants that were classified as non-CF-risk in *CFTR* (p-value=0.37 and 0.56, Table 3). Taken together, these results confirm our hypothesis that rare variants that impair *CFTR* function reduce the risk of IBD.

**Table 3.**
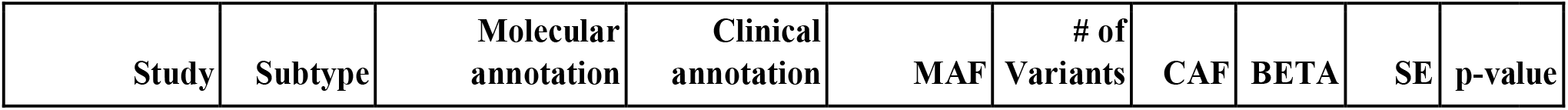
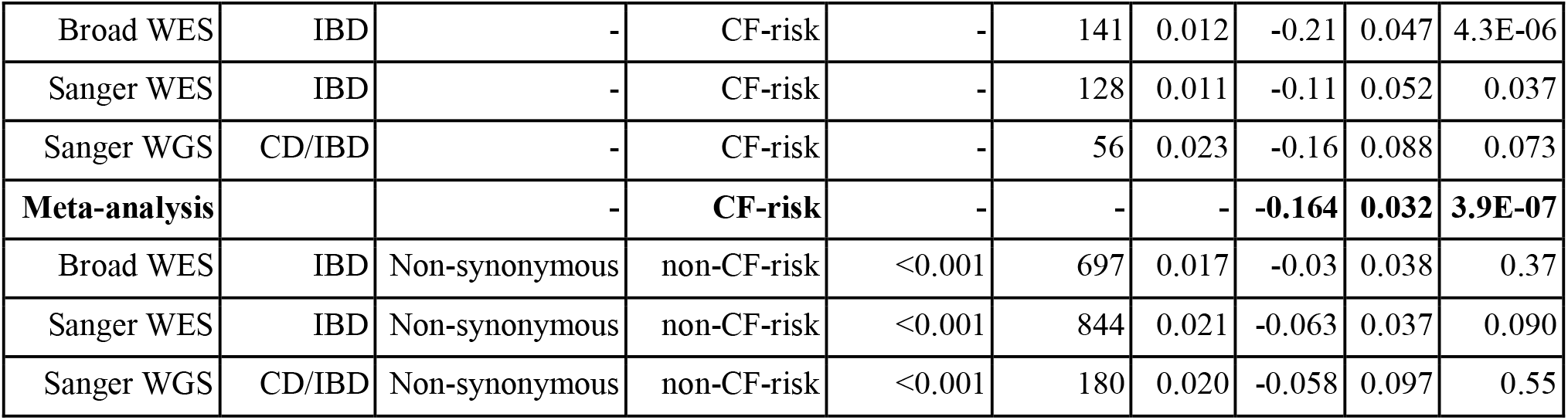
*CFTR* burden test results using clinical variant annotations. CAF: composite allele frequency, defined as the frequency of observing carriers of at least one variant of interest in the study. We applied an MAF upper bound to burden tests using non-CF-risk variants so that the CAFs are similar to those of CF-risk variants.

**Table 4.**
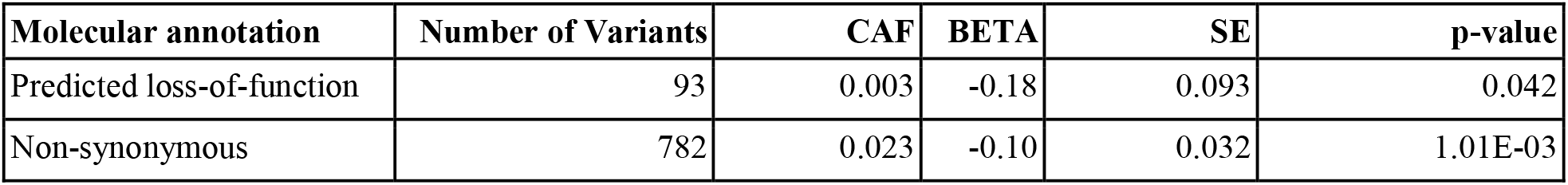
*CFTR* burden test using molecular annotations. Definitions of “predicted loss-of-function” and “non-synonymous” variants are described in Methods. We restricted the tests to variants with MAF<0.1%.

| Molecular annotation | Number of Variants | CAF | BETA | SE | p-value |
| --- | --- | --- | --- | --- | --- |
| Predicted loss-of-function | 93 | 0.003 | -0.18 | 0.093 | 0.042 |
| Non-synonymous | 782 | 0.023 | -0.10 | 0.032 | 1.01E-03 |

### Variant annotation plays a critical role in burden tests

The power of rare variant burden tests increases with the composite allele frequency and the proportion of causal to non-causal rare variants included in the analysis. Therefore, identifying a variant set that maximizes the inclusion of causal variants while minimizing non-causal variants is crucial to enhancing the power of the burden test. A common approach employed by the burden test is to use predicted loss of function variants or rare (MAF < 0.1%) non-synonymous variants. In both scenarios, we found much weaker evidence of association (p-value=1.01E-03 and 0.042, respectively, Table 3) compared to the burden test using CF-risk variants. This demonstrates that clinical variant annotations from the CFTR2 database outperform naive molecular consequence annotations in detecting the impact of rare *CFTR* variants on IBD risk.

Unfortunately, very few genes are as thoroughly studied and clinically annotated as *CFTR*. Most human genes lack accurate variant effect annotation. *In silico* variant pathogenicity predictions can thus play an important role in systematically annotating variants to enhance the power of rare variant burden tests. Recently, Alpha Missense (AM)^25^, a machine learning model trained on protein structure prediction and evolutionary constraint information for predicting missense variant pathogenicity, has been shown to have more accurate missense pathogenicity predictions as evidenced by better correlations with functional assays than previous prediction models^26^. Thus, we evaluated the utility of AM for prioritizing *CFTR* missense variants for inclusion in burden tests, using the CFTR2 database as the benchmark.

We annotated 698 *CFTR* missense variants available in the Broad WES dataset, among which 74 are CF-risk variants according to the CFTR2 database (Methods). We observed that most CF-risk variants were predicted as “likely pathogenic” (AM score>0.56) (Figure 2a), and variants with high AM scores (>0.8) had stronger protective effects against IBD (Figure 2b). However, 73% of predicted “likely pathogenic” variants were non-CF-risk in CFTR2. This finding is concordant with those from previous benchmarking studies that showed AM, as well as other pathogenicity prediction models, tend to overcall pathogenic variants^26,27^. The power to detect association via a variant burden test could therefore be heavily influenced by the AM score threshold used to define the set of variants included in the test. To investigate this, we performed burden tests by selecting *CFTR* missense variants (restricted to MAF < 0.1%) with AM score threshold ranging from 0 to 1 in increments of 0.01.

**Figure 2:**
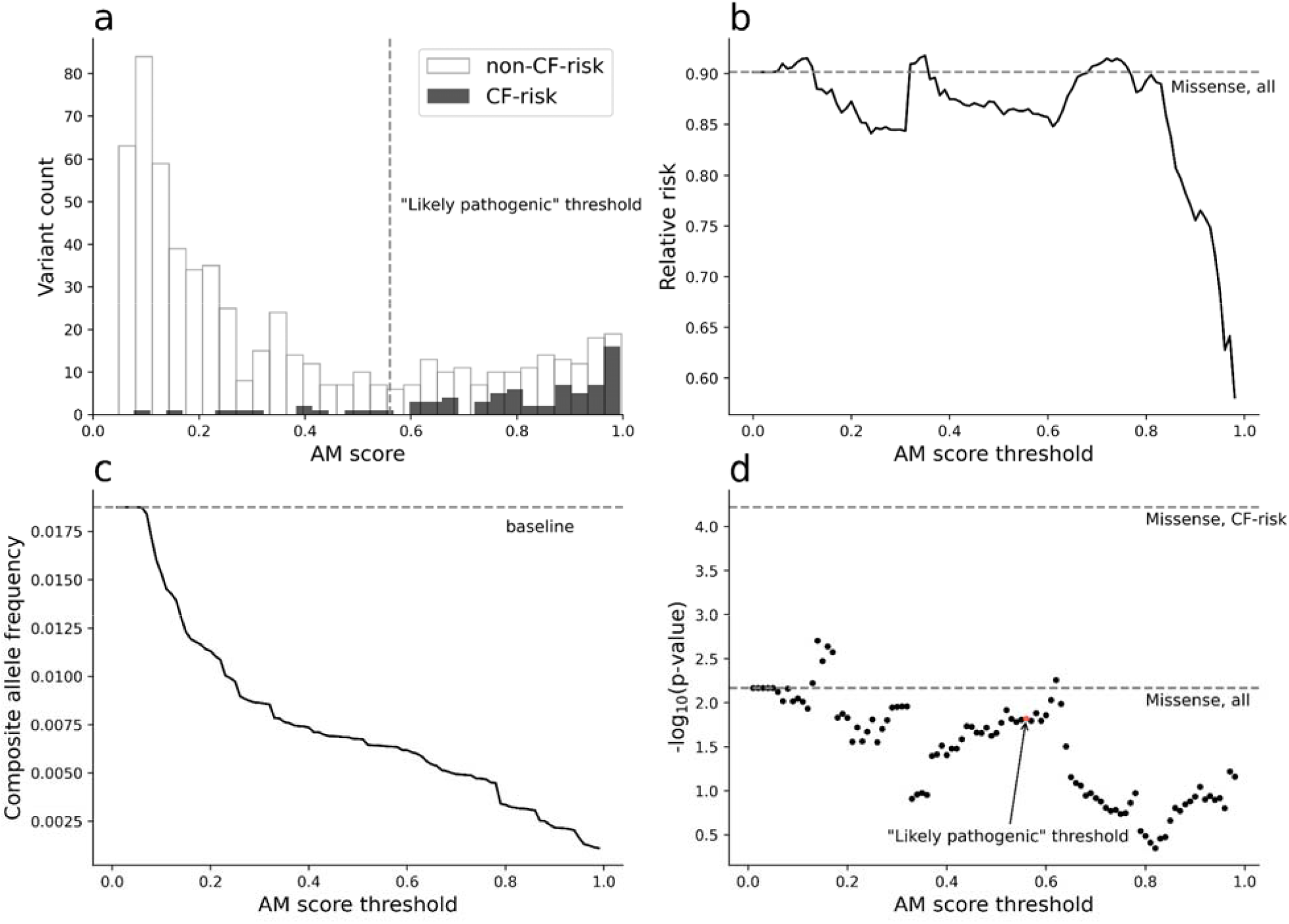
Prioritizing *CFTR* variants in burden tests using AlphaMissense. **a)** AM scores for CF-risk variants and non-CF-risk variants. **b)** Relative risk for missense variants with AM score above the AM threshold (x-axis). Relative risk was calculated as CAF in cases / CAF in controls. **c)** CAFs for missense variants with scores above the AM threshold (x-axis). **d)** Burden test significance using missense variants with AM scores above the threshold (x-axis). The result using the AM “likely pathogenic” threshold (0.56) is marked with red. We restricted the tests to variants with MAF<0.1%.

Our analysis showed that a higher AM score threshold does not always improve the statistical power of the burden test. Statistical power is sensitive to both the effect size (increases with the AM score threshold, Figure 2b) and the composite allele frequency (CAF, decreases with the AM score threshold, Figure 2c). Specifically, within the score range of 0.56 to 1, the burden test significance decreased with more stringent AM score cutoffs due to a reduction in CAF, despit a stronger effect size (Figure 2d). To effectively incorporate pathogenicity scores in burden tests, such as AM, further improvements on *in silico* variant annotations are needed to match the accuracy of clinical annotations (Discussion).

## Discussion

Leveraging a large-scale IBD exome sequencing dataset, we showed at single-variant and gene-based levels that CF-risk variants in the *CFTR* gene confer a protective effect against IBD. The gut epithelial barrier plays a fundamental role in maintaining intestinal homeostasis and protecting against IBD^28^. Given *CFTR* mutations are associated with mucus buildup on the epithelial cell surface, it is possible that heterozygous *CFTR* mutations alter the intestinal mucosa which provides an enhancing effect on the gut epithelial barrier. Using a CF mouse model, Gabriel *et al*^*29*^ showed that heterozygote carriers of pathogenic *CFTR* variants are resistant to cholera toxin. This resistance is due to reduced intestinal fluid and chloride ion secretion in response to the toxin. In addition, it has been demonstrated that *CFTR* can serve as an epithelial receptor for S. Typhi transluminal migration and that heterozygous deltaF508 mice translocated significantly fewer S. Typhi into the gastrointestinal submucosa than wild-type *CFTR* mice^30^. Therefore, it is also plausible that the protective effect of *CFTR* in IBD may stem from similar interactions with yet unidentified bacteria^19^. Our finding suggests a previously under-appreciated role of CF-risk mutations in human health and disease. Follow-up experimental studies are warranted to investigate the exact cellular pathways CF-risk mutations disrupt to exert such a protective effect. This may provide insights for an effective therapeutic intervention for IBD.

The ascertainment bias in CF patients can potentially confound this study. The Broad study ascertained control subjects from hospitals, while the Sanger study mostly ascertained controls from the community via UK Biobank. Therefore, an over-representation of CF patients in the Broad study can potentially create a spurious association between CF-risk variants and IBD. We controlled for this confounding factor by removing all predicted CF patients based on genetic data. While this approach is an approximation and does not guarantee the removal of all CF patients, the remaining number of CF patients is very small and should not affect the validity of our conclusion.

Burden tests are sensitive to the quality of variant annotations. While the clinical evidence-based *CFTR* variant annotations established the protective effect of CF-risk variants on IBD, simply including all predicted loss-of-function or missense variants in the burden test failed to reach exome-wide statistical significance. We evaluated AM, the best-in-class *in-silico* variant pathogenicity predictor, and found that its effectiveness in variant prioritization for burden tests is hampered by insufficient accuracy. Further improvement in pathogenicity prediction models for missense variants is needed. Furthermore, a higher score threshold for selecting variants for burden tests does not guarantee improvement in statistical power as it may sacrifice sensitivity for specificity, both of which are needed for a powerful burden test.

While this study focuses on the effect of *CFTR* mutations on IBD, the exome-wide analysis, incorporating additional samples sequenced at Sanger Institute, is underway. In the very near future, we expect to report a comprehensive list of IBD-associated genes through large-scale exome sequencing analysis.

## Data Availability

Genome Reference Consortium Human Build 38 can be accessed at https://www.ncbi.nlm.nih.gov/assembly/. Cystic Fibrosis variant annotation data can be downloaded from the Variant List History tab on CFTR2.org. AlphaMissense pathogenicity scores can be downloaded from https://zenodo.org/records/8208688. Sequence data used in this study has been made publicly available in dbGaP Study Accession: phs001642.v2.p1 - Center for Common Disease Genomics [CCDG] - Autoimmune: Inflammatory Bowel Disease (IBD) Exomes and Genomes (https://www.ncbi.nlm.nih.gov/projects/gap/cgi-bin/study.cgi?study_id=phs001642.v2.p1).

## ACKNOWLEDGMENTS

We thank all of the principal investigators, local staff from individual cohorts, and all of the patients who kindly donated samples used in the study for making possible this global collaboration and resource to advance IBD genetics research. This research was funded in whole, or in part, by the US National Institutes of Health grants no. U54HG003067 and no. 5UM1HG008895, the Wellcome Trust grants no. [206194] and no. [108413/A/15/D], and The Leona M. & Harry B. Helmsley Charitable Trust grant no. 2015PG-IBD001. We thank the Broad Institute Genomics Platform for genomic data generation efforts and the Stanley Center for Psychiatric Research at the Broad Institute for supporting control sample aggregation. We thank the Sanger Institute Scientific Operations teams and Human Genetics Informatics team for sample handling and data generation. This research was supported by the NIHR IBD BioResource and NIHR Biomedical Research Centres in Cambridge, Oxford, Imperial, UCH and Newcastle. The views expressed are those of the authors and not necessarily those of the NIHR or the Department of Health and Social Care. The NIHR IBD BioResource acknowledges co-funding by Crohn’s Colitis UK and The Leona M. and Harry B. Helmsley Charitable Trust. H.H. acknowledges support from NIDDK grant no. R01DK129364 and the Stanley Center for Psychiatric Research. Individual studies contributing to this project acknowledge support from NIH grants no. DK062431, no. DK062432, no. DK087694, no. K23DK117054, no. R01DK111843, no. P01DK094779, no. R01HG010140, no. 5U01HG009080 and no. DK062420, and NIDDK grants no. P01DK046763, no. U01DK062413, DK 043351 and no. R01DK104844.

## AUTHOR CONTRIBUTIONS

H.H., C.A.A. and M.J.D. designed and supervised the study. C.R.S., L.F., H.H., C.A.A. and M.J.D. were responsible for project management. M.Y., Q.Z., K.Y., and A.S. performed data analysis. M.Y. and H.H. wrote the manuscript. All authors have reviewed and approved the manuscript.

## METHODS

### Ethics

All relevant ethical guidelines have been followed, and any necessary institutional review board (IRB) and/or ethics committee approvals have been obtained. The Broad Institute component of this study was approved under Study Protocol 2013P002634 (the Broad Institute Study of Inflammatory Bowel Disease Genetics), and undergoes annual continuing review by the Mass General Brigham Human Research Committee IRB of Mass General Brigham (Mass General Brigham IRB). Approval was given on 27 January 2021 for this study. DNA samples sequenced at the Sanger Institute were ascertained under the following ethical approvals: 12/EE/0482, 12/YH/0172, 16/YH/0247, 09/H1204/30, 17/EE/0265, 16/WM/0152, 09/H0504/125, 15/EE/0286, 11/YH/0020, 09/H0717/4, REC 22/02, 03/5/012, 03/5/012, 2000/4/192, 05/Q1407/274, 05/Q0502/127, 08/ H0802/147, LREC/2002/6/18, GREC/03/0273 and YREC/P12/0. All informed consent from participants has been obtained, and the appropriate institutional forms have been archived.

### Broad Institute data production

#### Sequencing

The sequencing process included sample preparation (Illumina Nextera, Illumina TruSeq, and Kapa Hyperprep), hybrid capture (Illumina Rapid Capture Enrichment - 37 Mb target [“Nextera”, Table 1], and Twist Custom Capture - 37 Mb target [“Twist”, Table 1]) and sequencing (Illumina HiSeq2000, Illumina HiSeq2500, Illumina HiSeq4000, Illumina HiSeqX, Illumina NovaSeq 6000, 76-base pair (bp) and 150-bp paired reads). Sequencing was performed at a median depth of 85% targeted bases at >20×. Sequencing reads were mapped by BWA-MEM to the hg38 reference using the GATK ‘functional equivalence’ pipeline. The mapped reads were then marked for duplicates, and base quality scores were recalibrated. They were then converted to CRAMs using Picard 2.16.0-SNAPSHOT and GATK 4.0.11.0. The CRAMs were then further compressed using ref-blocking to generate gVCFs. These CRAMs and gVCFs were then used as inputs for joint calling. To perform joint calling, the single-sample gVCFs were hierarchically merged.

#### Quality control

Quality control (QC) analyses were conducted in Hail v.0.2.128 (Supplementary Figure 1). We first split multiallelic sites and coded genotypes with low genotype quality (GQ < 20) as missing. To exclude variant sites that fall outside of exome capture, we removed variants that are not annotated as frameshift, inframe deletion, inframe insertion, stop lost, stop gained, start lost, splice acceptor, splice donor, splice region, missense or synonymous. Sample-level QC. Samples that satisfy the following conditions were removed: (1) samples with an extremely large number of singletons (≥500); (2) samples with mean GQ < 30; (3) samples with missingness rates > 10%; (4) samples with outlying heterozygosity (±5 s.d. away from mean within the population); (5) samples with inconsistent genetically imputed sex and reported sex; and (6) duplicated samples, which were removed by identifying pairs of samples sharing identical genotypes (PI_HAT > 0.95) and keeping the sample with higher mean GQ. Variant-level QC. Variants that satisfy following conditions were removed: (1) variants with missingness rate > 5%; (2) variants with mean read depth (DP) < 10; (3) variants with >10% samples that were heterozygous and with an allelic balance ratio <0.3 or >0.7; and (4) variants that have known quality issues in both gnomAD v2 and v3 dataset (non-empty values in the “filter” column).

#### Ancestry assignment

We trained a random forest classifier using 1000 Genomes Project (1KGP) subjects and ten principal components (PC) derived from a set of ∼22000 common variants shared between 1KGP and our callset. We projected all our samples onto the PC space generated based on the 1KGP subjects and assigned each of our subject to European (CEU, TSI, FIN, GBR, IBS), African and American (YRI, LWK, GWD, MSL, ESN, ASW, ACB, MXL, PUR, CLM, PEL), East Asian (CHB, JPT, CHS, CDX, KHV), and South Asian (GIH, PJL, BEB, STU, ITU). For this study, we kept samples that were classified as European with a prediction probability greater than 80% (Supplementary Figure 2).

### Sanger Institute data production

#### Sequencing

Genome sequencing was performed at the Sanger Institute using the Illumina HiSeqX platform with a combination of PCR and PCR-free library preparation protocols. Sequencing was performed at a median depth of 18.6x. Exome sequencing of IBD cases was performed at the Sanger Institute using the Illumina NovaSeq 6000 and the Agilent SureSelect Human All Exon V5 capture set. Controls from the UK Biobank were sequenced separately as a part of the UKBB WES200K release using Illumina NovaSeq and the IDT xGen Exome Research Panel v1.0 capture set (including supplemental probes). 168,100 UKBB participants with EUR ancestry were selected for use as controls, excluding participants with recorded or self-reported CD, UC, unspecified noninfective gastroenteritis or colitis, any other immune-mediated disorders, or a history of being prescribed any drugs used to treat IBD. Reads were mapped to hg38 reference using BWA-MEM 0.7.17. Variant calls were performed using DeepVariant and saved as per-sample gVCFs. These gVCFs are aggregated with GLnexus into joint-genotyped, multi-sample project-level VCFs (pVCFs). Variant calling was limited to Agilent extended target regions. Per-region VCF shards were imported into the Hail software and combined. This study considered variants located in intersection regions of Agilent and IDT exome captures + 100bp buffer.

#### *Quality control* Exome sequencing

A combination of filters was used to identify low-quality variants and samples. Genotype calls with low genotype quality (GQ < 20) in rare variants (MAF < 0.1%) were set as missing. A variant level QC was then applied by keeping variants that meet all the following criteria in both case (Sanger WES) and control (UKBB WES) samples: (1) mean GQ > 30; (2) mean read depth (DP) > 10; and (3) call rate > 0.95. Samples satisfying any of the following conditions were removed: (1) low average GQ (≤ 30); (2) low call rate (≤ 0.9); (3) disagreement between genetically predicted and reported sex; (4) genetically identified duplicates (samples with higher call rate were retained); and (5) high rates of heterozygosity (± 4 s.d. away from mean within each dataset). Genome sequencing: we applied variant quality score recalibration (VQSR) to calculate the variant quality score log-odds (VQSLOD) for each variant using GATK v4.4. Variants in the range of VQSLODs corresponding to the remaining 0.5% of the truth set were removed. Furthermore, we kept variants that meet all the following criteria: (1) mean GQ > 30; (2) mean read depth (DP) > 10; and (3) call rate > 0.9. Details on sample QC were available elsewhere^22^.

#### Ancestry assignment

We selected a set of ∼14,000 high-quality common variants that were shared between our subjects and 1KGP subjects for ancestry assignment. Using this set of variants, we created four principal components from the 1KGP subjects and projected our subjects to these components. We then used Random Forest to classify samples into broad genetic ancestry groups (EUR, AFR, SAS, EAS, admixed), with 1KGP as the training dataset. We only retained the EUR samples for this study, as the number of cases for other ancestry groups was too small for robust association analysis.

### Association analysis. Broad Institute

Association analyses were performed using a logistic mixed model implemented in REGENIE v.2.2.4. A set of high-confidence variants (>1% MAF, 99% call rate, LD-pruned) was used for polygenic effect parameter estimation (Step 1). To control for case-control imbalance, Firth correction was applied to association tests with p-value < 0.05. To control for residual population structure, we calculated five PCs using a set of well-genotyped common SNPs, excluding regions with known long-range LD. We included a binary variable representing the sample sequencing platform to control for the sequencing heterogeneity (Twist or Nextera). These variables were used as covariates in both single-variant and burden tests, along with sex and the polygenic effect parameter calculated in Step 1. **Sanger Institute:** A similar pipeline has been applied at the Sanger Institute. We used REGENIE v3.1.2 to calculate the leave-one-chromosome-out (LOCO) score (Step 1) based on a set of high-quality variants (MAF > 1%, HWE P > 1e-15, LD-pruned with R2=0.9, and not from long-range LD regions). We then performed logistic regression using the LOCO score, sex, and four PCs as covariates in the model (Step 2). PCs were calculated based on variants with MAF > 0.1% and HWE P > 1e−15; variants from long-range LD regions or known IBD regions were excluded. We applied the fast Firth correction (flags: “--firth --approx”) to association tests with p-values < 0.05 to correct for effect size estimation bias caused by case-control imbalance.

### Meta-analysis

We used METAL^31^ with an IVW fixed-effect model to meta-analyze the association statistics across different studies.

### Clinical variant Annotation

Cystic Fibrosis variant annotation was downloaded from the Variant List History tab on CFTR2.org on April 23rd, 2024. Each variant was mapped to variant ID in GRCh38 by its cDNA name.

### Variant annotation for molecular effect

Non-synonymous variants were classified using Ensembl Variant Effect Predictor (VEP v.95.0 in Broad institute and VEP v.110.1 in Sanger institute)^32^ as one of the following most severe consequences: “frameshift_variant”, “stop_gained”, “splice_acceptor_variant”, “splice_donor_variant”, “inframe_deletion”, “inframe_insertion”, “stop_lost”, “start_lost”, “missense_variant”. Predicted loss-of-function variants are defined as variants annotated by VEP as one of the following mutation types: “frameshift_variant”, “stop_gained”, “splice_acceptor_variant”, “splice_donor_variant”.

### Variant Annotation using AlphaMissense

AlphaMissense (AM) predictions for all single amino acid substitutions in the human proteome data were downloaded and subsetted to 698 CFTR missense variants available in the Broad discovery dataset.

## CODE AVAILABILITY

The software and code used are described throughout the Methods and can be found at https://github.com/mingRYU/CFTR-IBD.

