## Supplementary Figures 1-2 for "Cystic fibrosis risk variants confer protection against inflammatory bowel disease"

| **Supplementary Figure** | **Title** |
| --- | --- |
| 1 | Quality control workflow |
| 2 | Population assignment with principal components |

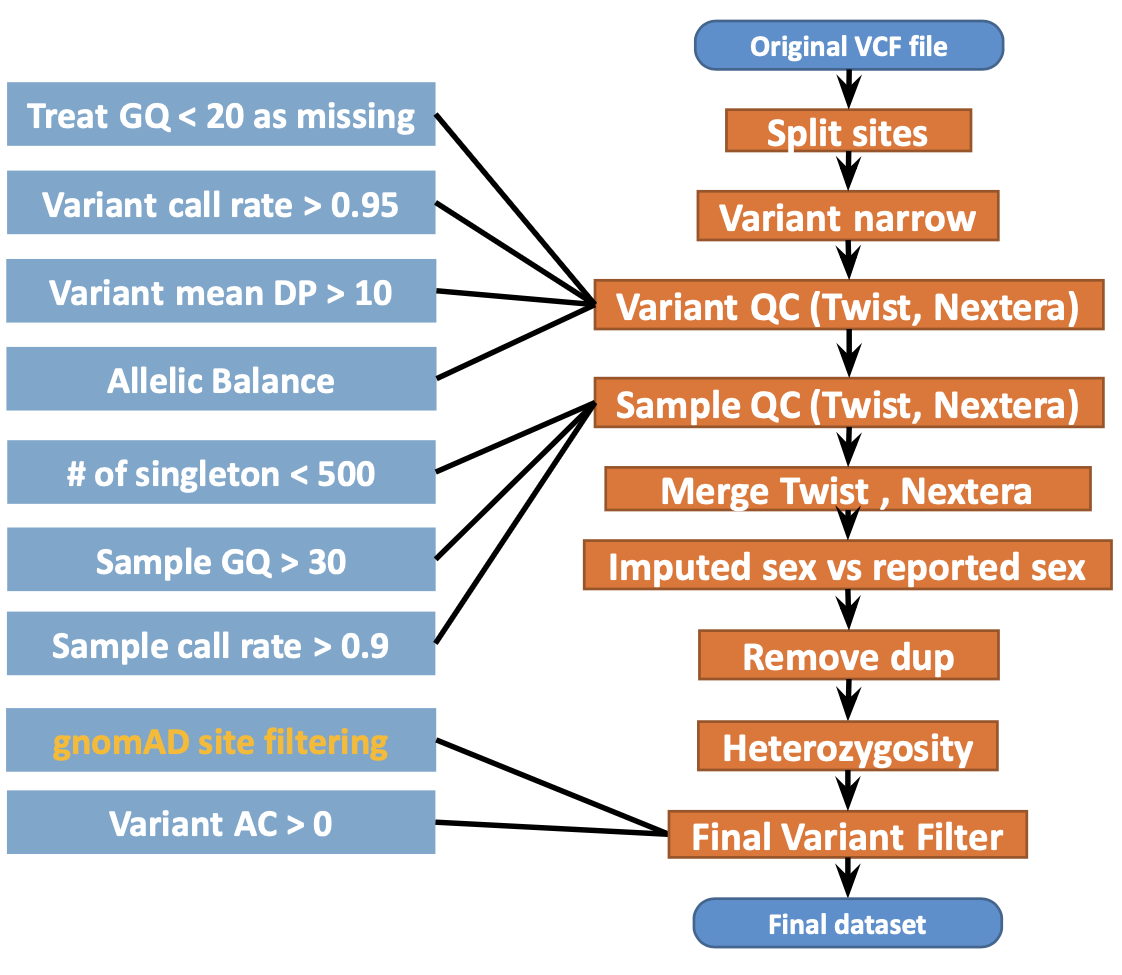
**Supplementary Figure 1: Quality control workflow.** We show the quality control steps performed on variants and subjects on the Broad sequencing dataset. Details and specific parameters are described in Methods.

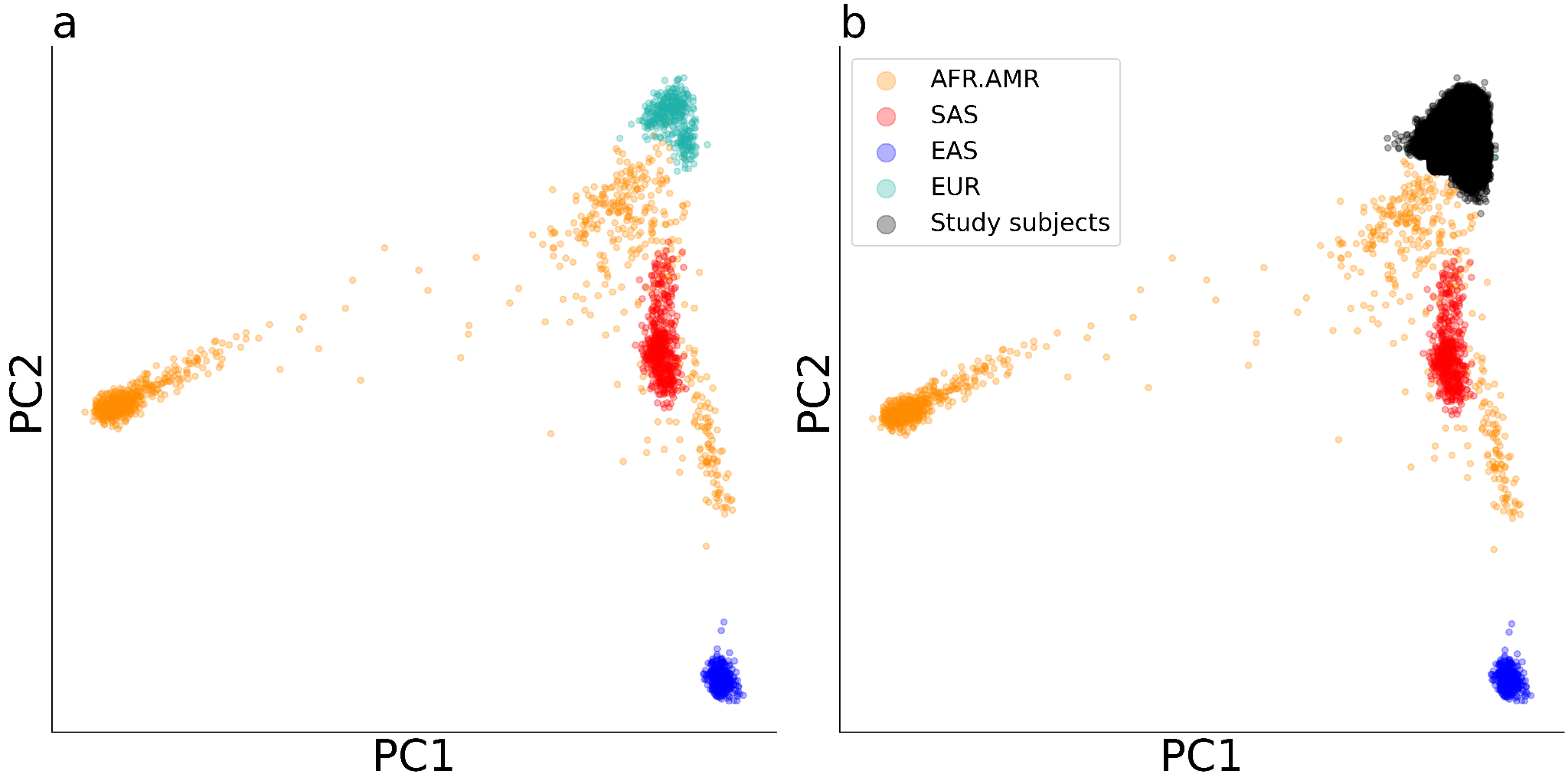

**Supplementary Figure 2: Population assignment with principal components.** Principal components (PCs) were calculated for the 1000 Genomes Project Phase III subjects. Genetic ancestries are indicated by their colors. **a)** subjects from the 1000 Genomes Project. **b)** Study subjects from the Broad Institute were projected to the same PC space. Black dots represent study subjects of European ancestries that entered into the analysis.
